# Mental disorders in adolescents at familial high-risk of schizophrenia or bipolar disorder and population-based controls – an eight-year follow-up study, The Danish High Risk and Resilience Study, VIA 15

**DOI:** 10.64898/2026.08.22.26360313

**Authors:** Doris Helena Bjarnadóttir Streymá, Maja Gregersen, Nanna Weye, Carsten Hjorthøj, Mette Falkenberg Krantz, Anne Søndergaard, Marta Schiavon, Sinnika Birkehøj Rohd, Martin Wilms, Ditte Ellersgaard, Sofie Baltser Christensen, Mette Enevoldsen, Merete Birk, Charlotte Sand Nielsen, Andreas Færgemand Laursen, Anette Faurskov Bundgaard, Lotte Veddum, Ole Mors, Aja Neergaard Greve, Nicoline Hemager, Merete Nordentoft, Anne AE Thorup

**Affiliations:** CORE-Copenhagen Research Center for Mental Health, Copenhagen University Hospital, Mental Health Center Copenhagen, Capital Region of Denmark, Copenhagen, Denmark; University of Copenhagen – Faculty of Health and Medical Sciences, Institute of Clinical Medicine, Copenhagen, Denmark; Research Unit at Child and Adolescent Mental Health Center, Copenhagen University Hospital, Mental Health Services, Capital Region of Denmark, Copenhagen, Denmark; Department of Public Health, Section of Epidemiology University of Copenhagen, Copenhagen, Denmark; Department of Clinical Medicine, Faculty of Health and Medical Sciences, Aarhus University, Aarhus Denmark; Psychosis Research Unit, Aarhus University Hospital Psychiatry, Aarhus, Denmark

## Abstract

**Background:** Children of parents with schizophrenia (SZ) or bipolar disorder (BP) show elevated rates of mental disorders. Longitudinal studies comparing offspring at familial risk with the background population are lacking.

**Method:** This study is an eight-year follow-up of the Danish High Risk and Resilience study. We examined four-year prevalence from age 11 to age15 (n=416), cumulative incidence by age 15 (n=516), persistency of mental disorders from age 11to age 15 (n=396) and global functioning in 15-year-old adolescents with familial high risk of SZ (FHR-SZ) or BP (FHR-BP) compared to population-based controls (PBC). We assessed mental disorders and global functioning with the Kiddie Schedule for Affective Disorders and Schizophrenia – Present and Lifetime Version (K-SADS-PL) and the Children’s Global Assessment Scale (CGAS).

**Results:** Four-year prevalence of any mental disorder was higher in FHR-SZ (51.3%, OR=2.39, 95% CI 1.49-3.83) and FHR-BP (45.9%, OR=1.98, 95% CI 1.16-3.37) compared with PBC (30.5%). Cumulative incidence of mental disorders by age 15 was higher in FHR-SZ (67.2%, OR=3.19, 95% CI 2.11-4.82) and FHR-BP (64.4%, OR=2.82, 95% CI 1.75-4.54) than in PBC (39.1%). Adolescents with FHR-SZ showed the highest rate of persistent mental disorders (33.3%), followed by FHR-BP (24.5%), and PBC the lowest (12.9%). Global functioning at age 15 was lower in FHR-SZ than in both FHR-BP and PBC, and FHR-BP showed lower scores compared with PBC. Between-group differences in cumulative incidences of mental disorders and in global functioning scores remained stable across ages 7,11 and 15.

**Conclusion:** Adolescents at FHR-SZ or FHR-BP show elevated risks of a range of mental disorders, psychiatric comorbidity, and lower global functioning from childhood to mid-adolescence, not confined to the disorders for which they carry familial risk. This vulnerability underscores the need for early detection and support for FHR offspring and their families.

**Key Point:** *What’s known?:*

- Children of parents with schizophrenia or bipolar disorder (Familial High Risk (FHR) offspring) have an elevated risk of mental disorders. Adolescence is a recognized critical period for mental disorder onset and progression.

*What’s new?:*

- This eight-year prospective follow-up study brings novel insight into the transition from childhood to mid-adolescence, by comparing FHR offspring with population-based controls.
- Our study showed higher four-year prevalence, cumulative incidence and persistency of mental disorders as well as lower global functioning in the FHR.
- Adolescent psychopathology encompassed a broad range of diagnoses beyond parental disorders.

*What’s relevant?:*

- Group differences in mental disorders and global functioning remained stable from childhood to adolescence, indicating a persistent, early emerging vulnerability, which underscores the importance of early identification and family support.

## Introduction

Schizophrenia (SZ) and bipolar disorder (BP) are severe mental disorders which cluster in families, reflecting both genetic and environmental contributions to their transmission across generations (Robinson et al., 2024; Robinson & Bergen, 2021).

Children of parents diagnosed with these disorders are susceptible to developing the same disorder as their parents and at increased risk of developing other mental disorders as well (Uher et al., 2023). This increased prevalence of mental disorders is evident already in early childhood (Davidsen et al., 2022).

However, not all children at familial high risk (FHR) will develop a mental disorder and FHR studies play an important role in examining the development of psychopathology. By identifying indicators of psychiatric vulnerability prior to the potential onset of mental disorder, these studies offer insights to guide preventive intervention strategies.

Previous studies of offspring at familial risk of SZ have consistently shown a higher prevalence of a broad spectrum of Axis I mental disorders, including attention-deficit/hyperactivity disorder (ADHD), disruptive behavior disorders, mood disorders, and anxiety disorders(Hans, Auerbach, Styr, & Marcus, 2004; Keshavan et al., 2008; Ross & Compagnon, 2001; Shah et al., 2019). Similarly, studies of offspring with familial risk of BP have reported elevated rates of these disorders (Maziade et al., 2008; Mesman, Nolen, Reichart, Wals, & Hillegers, 2013). For example, in the Dutch Bipolar Offspring Study, after 12 years of follow-up (N = 108) 72% of BP offspring had developed a lifetime Axis I disorder, 54% a mood disorder, and 13% bipolar disorder(Mesman et al., 2013). By 22-years follow-up, 80% met the criteria for any lifetime Axis I disorders, and 65% had developed a mood disorder, mostly with comorbidity (75%) (Helmink, Mesman, & Hillegers, 2024).

Although SZ and BP are distinct disorders, they share some clinical features and exhibit genetic overlap, which may contribute to overlapping patterns of psychopathology in offspring of parents with SZ and BP (Anttila et al., 2018; Uher et al., 2023). Only few studies have examined mental disorders in both groups of children at FHR(De la Serna et al., 2021, 2025; Erlenmeyer-Kimling & Cornblatt, 1987; Maziade et al., 2008; Sanchez-Gistau et al., 2015; Setiaman, Mesman, van Haren, & Hillegers, 2024). Most of them include children across various age groups or lack control groups, which limit robustness of the findings. The BASYS Study investigated offspring at FHR of SZ and BP together with a control group (N=238) and found elevated rates of Axis I disorders in offspring with FHR of SZ and offspring of FHR of BP compared to controls(Sanchez-Gistau et al., 2015). At baseline, two-year and four-year follow up, mood disorders were most prevalent in BP offspring, while SZ offspring had higher rates of ADHD, disruptive behavior and prodromal symptoms of psychosis compared to controls (De la Serna et al., 2021, 2025). Global functioning was lowest in SZ offspring compared with both BP offspring and controls (De la Serna et al., 2021).

In the Danish High Risk and Resilience Study (the VIA cohort), we have previously demonstrated an increased prevalence of mental disorders and reduced levels of global functioning in both children at FHR of SZ (FHR-SZ) and FHR of BP (FHR-BP), which persisted across childhood from age 7 to age 11(Ellersgaard et al., 2018; Gregersen et al., 2022). We have also demonstrated that children in the FHR-SZ group were less likely to recover from mental illness during childhood (Gregersen et al., 2022). Since adolescence is a critical period for the development of mental disorders, characterized by rapid hormonal changes, brain maturation, and heightened social and existential challenges, all of which can contribute to the onset of psychiatric symptoms, further research is required(Maughan & Collishaw, 2015; Solmi et al., 2022). To the best of our knowledge however, no existing study has examined the development of psychopathology from childhood to adolescence in a longitudinal cohort of offspring at FHR-SZ or FHR-BP with a very narrow age-range.

### Aim

By comparing 15-year-old adolescents at FHR-SZ or FHR-BP with population-based controls (PBC) in a clinical, longitudinal cohort, we aimed to investigate:

1. The four-year prevalence of mental disorders and psychiatric comorbidity from ages 11-15
2. The cumulative incidence of mental disorders and psychiatric comorbidity by age 15
3. Disorder persistency, defined as proportions of incident, remittent, persistent, and no mental disorders from ages 11-15
4. The global level of functioning at age 15 and the trajectories hereof from ages 7-15

## Methods

### Study population

The Danish High Risk and Resilience Study (the VIA cohort) is a nationwide, representative cohort study examining children born to parents diagnosed with SZ, BP, or neither of these conditions (i.e. PBC) (Krantz et al., 2023; Thorup et al., 2015, 2018, 2022). All participants were initially recruited from the Danish national registers. Parental diagnostic information was obtained from The Danish Psychiatric Central Research Register while data on children was obtained from The Danish Civil Registration System (Mors, Perto, & Mortensen, 2011; Pedersen, 2011). The study population included children born between September 1, 2004, and August 31, 2009, who had at least one parent diagnosed in the secondary healthcare sector with a schizophrenia spectrum disorder (ICD-10: F20, F22, F25; or ICD-8: 295, 297, 298.29, 298.39, 298.99) or bipolar disorder (ICD-10: F30, F31; or ICD-8: 296.19, 296.39), along with population-based control children. Children in the PBC group were matched by age, sex, and municipality to the FHR-SZ group. Although the FHR-BP children were not matched, they were comparable in terms of age and sex to the FHR-SZ and PBC group. At baseline, the children were assessed at age 7 (the VIA 7 study, N=522; FHR-SZ: 202, FHR-BP: 120; PBC: 200) and at four-year follow-up at age 11 (the VIA 11 study, N=465, FHR-SZ: 179, FHR-BP:105, PBC: 181), achieving an overall retention rate of 89%. The eight-year follow-up took place at age 15 (the VIA 15 study, N=427, FHR-SZ:158, FHR-BP:100, PBC:169) with an overall retention rate of 82%. The VIA cohort comprises a total of 16 sibling pairs. At each assessment, the primary caregiver, in most cases defined as the parent/adult who lived with and knew the adolescent best or spent the most time with the child/adolescent, contributed with information about the adolescent.

### Ethical considerations

The Danish Data Protection Agency approved the study, and the Danish Committee on Health Research Ethics concluded that ethical approval was unnecessary due to the study’s observational nature in the VIA 7 study, while ethical permission was obtained for the VIA 11 study (Protocol number H-16043682) and the VIA 15 study (Protocol number: H-20067908) (Thorup et al., 2015, 2018, 2022). Following a thorough explanation of the procedures, the legal guardians provided written informed consent for their own and their child’s participation, while adolescents gave oral consent (Thorup et al., 2022).

### Measures

Mental disorders were assessed using the gold-standard semi-structured interview, the Kiddie Schedule for Affective Disorders and Schizophrenia for School-Age Children—Present and Lifetime Version (K-SADS-PL), in all three VIA studies: the VIA 7 study, covering the period from birth to age 7; the VIA 11 study, covering ages 7 to 11; and the current VIA 15 study, focusing on ages 11 to 15 (Kaufman et al., 1997). Psychologists, research nurses, and medical doctors, who had received formal training in using the K-SADS-PL and were blinded to FHR status, conducted face-to-face interviews with the adolescent and/or their primary caregiver separately to assess mental disorders occurring since the previous assessment. Diagnoses were based on both DSM-IV and DSM-V criteria and confirmed at clinical conferences with a clinical professor, specialist in child and adolescent psychiatry (last author AAET), who was also blinded to FHR status. Following the K-SADS-PL all relevant and available information regarding the adolescent was considered to establish the most accurate diagnosis. As in our previous studies, elimination disorders, transient and unspecified tics, and specific phobias were excluded from the analyses owing to their uncertain clinical relevance (Ellersgaard et al., 2018; Gregersen et al., 2022). Psychiatric comorbidity was defined as having ≥2 mental disorders diagnosed at the same assessment.

Global functioning of the previous 30 days was assessed using the Children’s Global Assessment Scale (CGAS) (Shaffer et al., 1983). Functioning is scored on a scale between 1 and 100, and higher scores reflect better functioning. The final score was determined following the evaluation of current mental disorders at a clinical conference with the last author (AAET).

General intelligence was measured using the Reynolds Intellectual Screening Test (RIST) at the four-year follow-up (age 11)(Reynolds, & Kamphaus, 2003). The level of functioning of the primary caregiver was assessed using the Personal and Social Performance scale (PSP) which is a semi-structured interview. The PSP scale ranges from 1 to 100 with higher scores indicating higher levels of functioning during the previous 30 days (Morosini, Magliano, Brambilla, Ugolini, & Pioli, 2000). Information on the primary caregiver’s employment status, single-caregiver status, educational level, and the adolescent’s potential out-of-home placement was obtained through an anamnestic interview with the primary caregiver at the eight-year follow-up.

### Statistical analyses

The between-group differences in background characteristics were analyzed using either one-way analysis of variance (ANOVA) or chi-square test, followed by pair-wise comparisons in the event of a significant main effect of group. Four-year prevalence of mental disorders from ages 11-15 was reported for adolescents who participated in a K-SADS-PL interview by age 15. Group differences were assessed using logistic regression adjusted for sex and clustered on sibling identifier. Cumulative incidence of mental disorders by age 15 was assessed for all children with at least one valid K-SADS-PL interview at age 7, 11, or 15. We applied a rule-based imputation approach informed by the observed diagnoses at previous assessments for participants that had not participated in all three assessments. Specifically, if a participant was classified as having a mental disorder at an assessment, they were classified as having that condition across any unobserved subsequent assessment. Conversely, if a participant was not classified as having a mental disorder at previous assessments, we assumed the absence of the condition at unobserved subsequent assessments. Group differences in cumulative incidence by age 15 was assessed using logistic regression, adjusted for sex and clustered on sibling identifier, while differences in changes in cumulative incidence from age 7 to age 11 and 15 were evaluated using generalized estimating equations logistic regression models with FHR, time, sex and time-by-FHR group interaction as covariates. Sibling identifier was specified as the clustering variable and an unstructured correlation matrix was used to model within-subject correlations flexibly. We used Wald test to assess diverging trajectories of cumulative incidence of mental disorders between FHR groups.

Disorder persistency was assessed by examining whether participants met criteria for any mental disorder occurring within the past four years at two time points: age 11 and age 15. Only participants with valid K-SADS-PL assessments at both time points were included. Changes in disorder status between these assessments were used to classify participants into four categories: (1) persistent disorder (any mental disorder present at both assessments), (2) incident disorder (any mental disorder absent at age 11 but present at age 15), (3) remittent disorder (any mental disorder present at age 11 but absent at age 15), and (4) no disorder (any mental disorder absent at both assessments). Differences in the distribution of these categories across all three study groups were examined using a Chi-square test.

Estimated marginal means of global functioning (CGAS scores) at age 15 and the trajectories of global functioning from ages 7 to age 11 and 15 were examined using linear mixed-effects models incorporating FHR group, time, child’s sex, and the time-by-group interaction as fixed factors and sibling identifier as random effect. We obtained estimated marginal means for each group at each assessment based on the model and used likelihood ratio test to assess differences in trajectories of global functioning.

Differences between participants and non-participants in the K-SADS-PL interview at age 15 were assessed using t-test for CGAS and chi-square tests for sex, FHR group, and any Axis I disorder measured at baseline.

An alpha threshold of <.05 was applied, with all p-values assessed using two-tailed tests. All analyses were performed in SPSS version 29.0.1.0 and R version 4.4.1.

We chose not to adjust for socioeconomic factors, as we considered these too closely related to the course of mental disorder.

## Results

### Background characteristics at age 15

A total of 416 adolescents participated in the K-SADS-PL interview (FHR-SZ, n=154; FHR-BP, n=98; PBC, n=164) at age 15 (table 1). The participating adolescents in the three groups were not significantly different in terms of sex and age at inclusion. Adolescents at FHR-SZ showed significantly lower scores of general intelligence, as assessed by RIST, at age 11 than the PBC group (p=0.03), whereas no significant difference was found between the adolescents at FHR-BP and the PBC group. Non-participants did not differ from participants at age 15 in terms of sex, FHR group, history of any Axis I mental disorder or level of functioning at age 7 (Table S1).

**Table 1:**
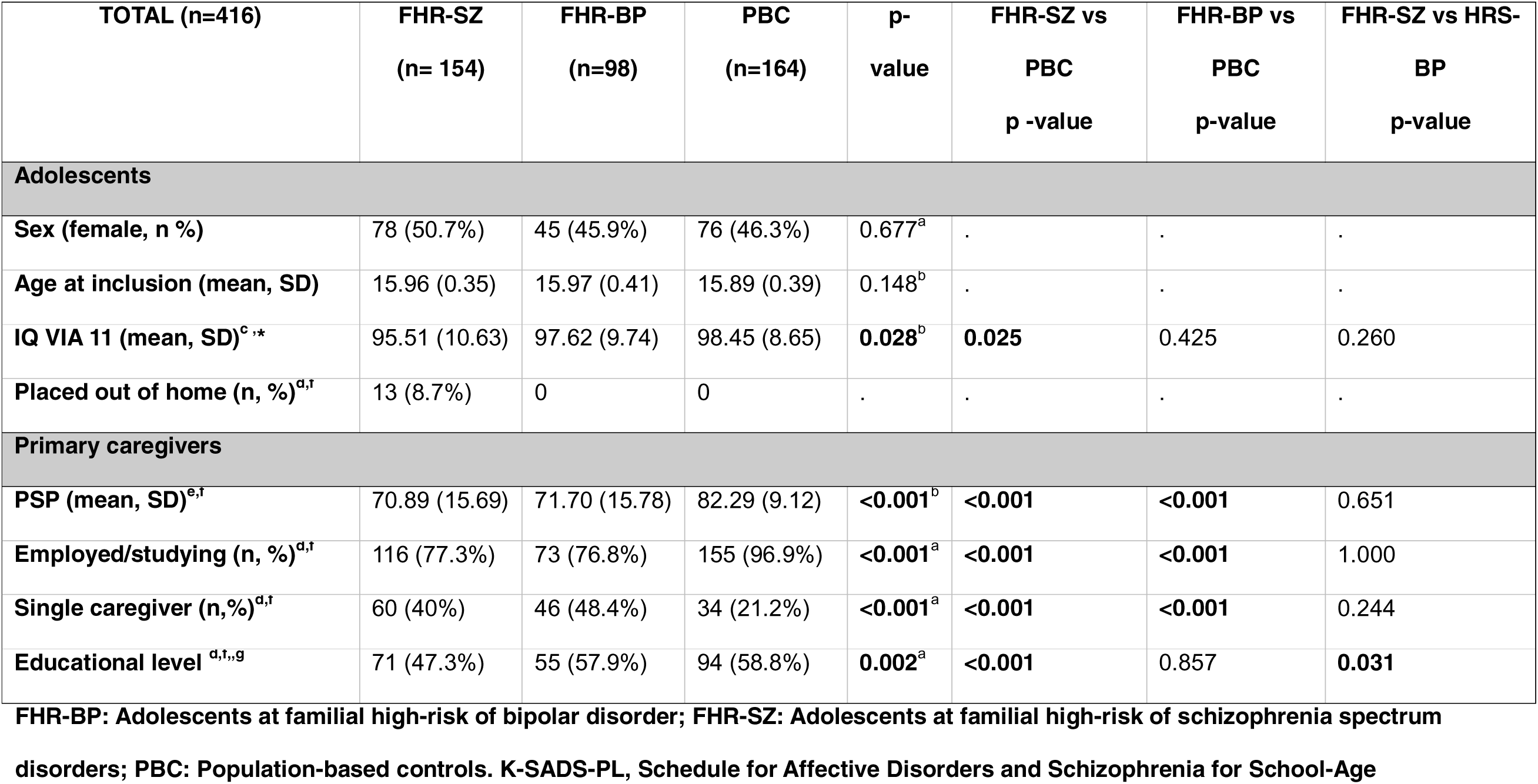
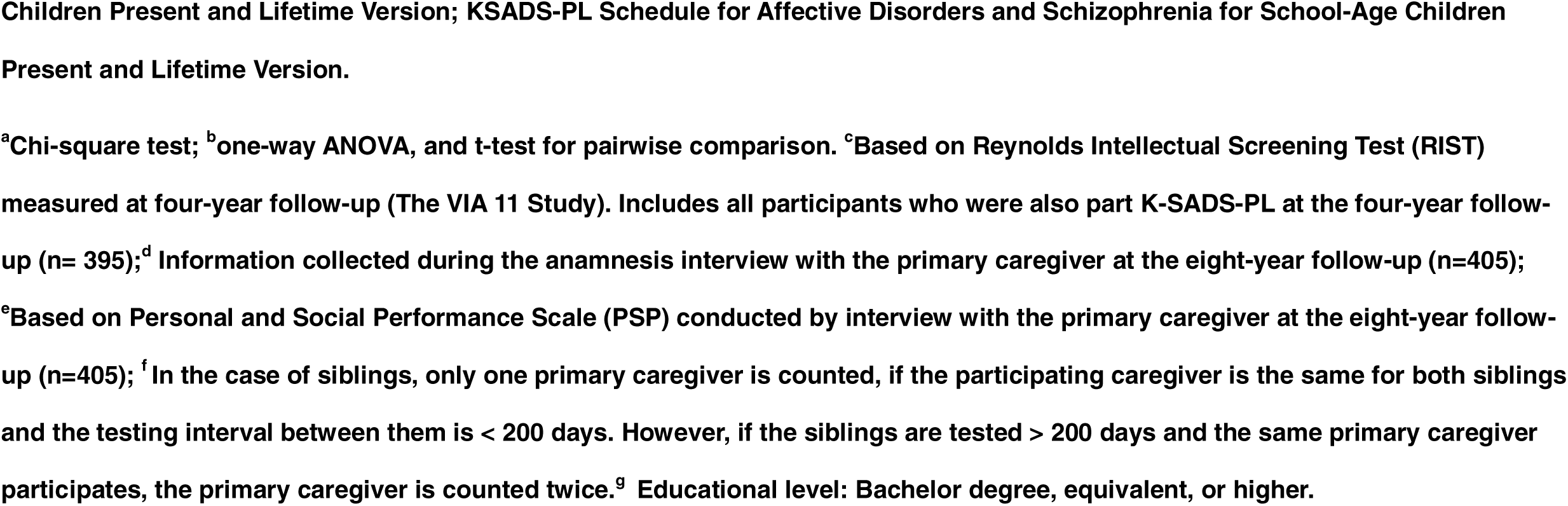
Background characteristics of 416 15-year-old adolescents and their primary caregivers in The Danish High Risk and Resilience Study at the eight-year follow-up (The VIA 15 Study).

Compared to the PBC group (97%), primary caregivers were significantly less likely to be employed or studying in the FHR-SZ group (77.3%, p<0.01) and the FHR-BP (76.8%, p<0.01). Additionally, significantly more caregivers were single caregivers in the FHR groups compared with the PBC group (40% among FHR-SZ, 48% among FHR-BP, and 21% among PBC (p<0.001)). Notably, out-of-home placement occurred exclusively among adolescents in the FHR-SZ group (n = 13, 8.7%) (Table 1).

### Four-year prevalence of mental disorders from age 11 to 15

Adolescents in both FHR groups had significantly higher prevalences of any Axis I mental disorder by age 15 compared to the PBC group. Furthermore, both FHR groups exhibited a higher prevalence of psychiatric comorbidity. The adolescents at FHR-SZ and FHR-BP exhibited three- and twofold higher odds of ADHD compared to the PBC group. Adolescents at FHR-SZ had the highest prevalence of autism spectrum disorders (14.9%), while adolescents in the FHR-BP group showed the highest prevalence of affective disorders (17.3%) and adjustment disorders (14.3%). Among the affective disorders, no cases of mania or hypomania were identified. Only adolescents in the FHR groups were diagnosed with psychotic disorders and posttraumatic stress disorder (PTSD), but too few were diagnosed to calculate differences across the groups (Table 2).

**Table 2:** Four-year prevalence of DSM-IV and DSM-V Axis 1 mental disorders and psychiatric comorbidity in 416 15-year-old adolescents in The Danish High Risk and Resilience Study assessed by the K-SADS-PL at the eight-year follow-up (the VIA 15 study). FHR-BP: Adolescents at familial high-risk of bipolar disorder; FHR-SZ: Adolescents at familial high-risk of schizophrenia spectrum disorders; PBC: Population-based controls. K-SADS-PL, Schedule for Affective Disorders and Schizophrenia for School-Age Children, Present and Lifetime Version. ^a^ Any Axis I disorder: Elimination disorders, transient and unspecified tics, and specific phobias were excluded; ^b^ Too few cases to calculate pair-wise comparison; ^c^ Tic disorders include Tourette’s disorder and chronic tic disorder; ^d^ Psychiatric comorbidity: The adolescent met the criteria for two or more different disorder categories at the eight-year follow up (between age 11 to 15).

|  | **FHR-SZ**<br>**(n= 154)** | **FHR-BP**<br>**(n= 98)** | **PBC**<br>**(n= 164)** | **FHR-SZ vs**<br>**PBC**<br>**(OR, 95% CI)** | **p-value** | **FHR-BP vs**<br>**PBC**<br>**(OR, 95% CI)** | **p-**<br>**value** | **FHR-SZ vs FHR-**<br>**BP**<br>**(OR, 95% CI)** | **p-**<br>**value** |
| --- | --- | --- | --- | --- | --- | --- | --- | --- | --- |
| **Any Axs 1 disorder<sup>a</sup>** | 79<br>(51.3%) | 45<br>(45.9%) | 50 (30.5%) | **2.39 (1.49-3.83)** | **<0.001** | **1.98 (1.16-**<br>**3.37)** | **0.013** | 1.21 (0.72-2.06) | 0.469 |
| **Affective Disorders** | 21<br>(13.6%) | 17<br>(17.3%) | 15 (9.1%) | 1.52 (0.75-3.15) | 0.242 | 2.12 (1.00-4.56) | 0.05 | 0.71 (0.35-1.46) | 0.346 |
| **Psychotic Disorders<sup>b</sup>** | < 5 | < 5 | 0 |  |  |  |  |  |  |
| **Anxiety Disorders** | 18<br>(11.7%) | 12<br>(12.2%) | 10 (6.1%) | 1.97 (0.88-4.65) | 0.107 | 2.25 (0.91-5.67) | 0.081 | 0.87 (0.39-1.99) | 0.745 |
| **ADHD** | 30<br>(19.5%) | 15<br>(15.3%) | 10 (6.1%) | **3.79 (1.78-8.08)** | **0.001** | **2.79 (1.20-**<br>**6.47)** | **0.017** | 1.37 (0.69-2.71) | 0.365 |
| **Disruptive Behaviour**<br>**Disorders<sup>b</sup>** | 7 (4.5%) | < 5 | < 5 |  |  |  |  |  |  |
| **Autism Spectrum**<br>**Disorders** | 23<br>(14.9%) | 13<br>(13.3%) | 11 (6.7%) | **2.44 (1.17-5.38)** | **0.02** | 2.13 (0.91-5.05) | 0.08 | 1.15 (0.56-2.46) | 0.712 |
| **PTSD<sup>b</sup>** | 5 (3.2%) | < 5 | 0 |  |  |  |  |  |  |
| **Adjustment Disorders** | 12 (7.8%) | 14<br>(14.3%) | 11 (6.7%) | 1.11 (0.47-2.67) | 0.814 | **2.42 (1.04-**<br>**5.81)** | **0.038** | 0.47 (0.20-1.08) | 0.096 |
| **Tic Disorders<sup>b,c</sup>** | 5 (3.2%) | < 5 | 9 (5.5%) |  |  |  |  |  |  |
| **Eating Disorders<sup>b</sup>** | 5 (3.2%) | 6 (6.1%) | < 5 |  |  |  |  |  |  |
| **Substance Use Disorders<sup>b</sup>** | < 5 | 0 | < 5 |  |  |  |  |  |  |
| **Psychiatric comorbidity<sup>d</sup>** | 31<br>(20.1%) | 26<br>(26.5%) | 17 (10.4%) | **2.14 (1.12-4.08)** | **0.021** | **3.19 (1.62-6.25)** | **0.001** | 0.67 (0.37-1.24) | 0.203 |

### Cumulative incidence of mental disorders by 15

A total of 516 children underwent at least one psychiatric assessment using the K-SADS-PL interview at age 7, 11, or 15, and a total of 44 children participated in one assessment, 78 in two assessments, and 394 participated in all three assessments. In total, 67.2 % of adolescents at FHR-SZ, 64.4% of adolescents at FHR-BP, and 39.1% of adolescents in the PBC group fulfilled the criteria for any Axis I mental disorder at any timepoint up until age 15 (Table 3). Psychiatric comorbidities were observed in 39% of adolescents at FHR-BP and in 31.3% of adolescents at FHR-SZ, corresponding to fourfold and threefold higher odds, respectively, compared with the PBC group (13.2%) (Table 3).

**Table 3:**
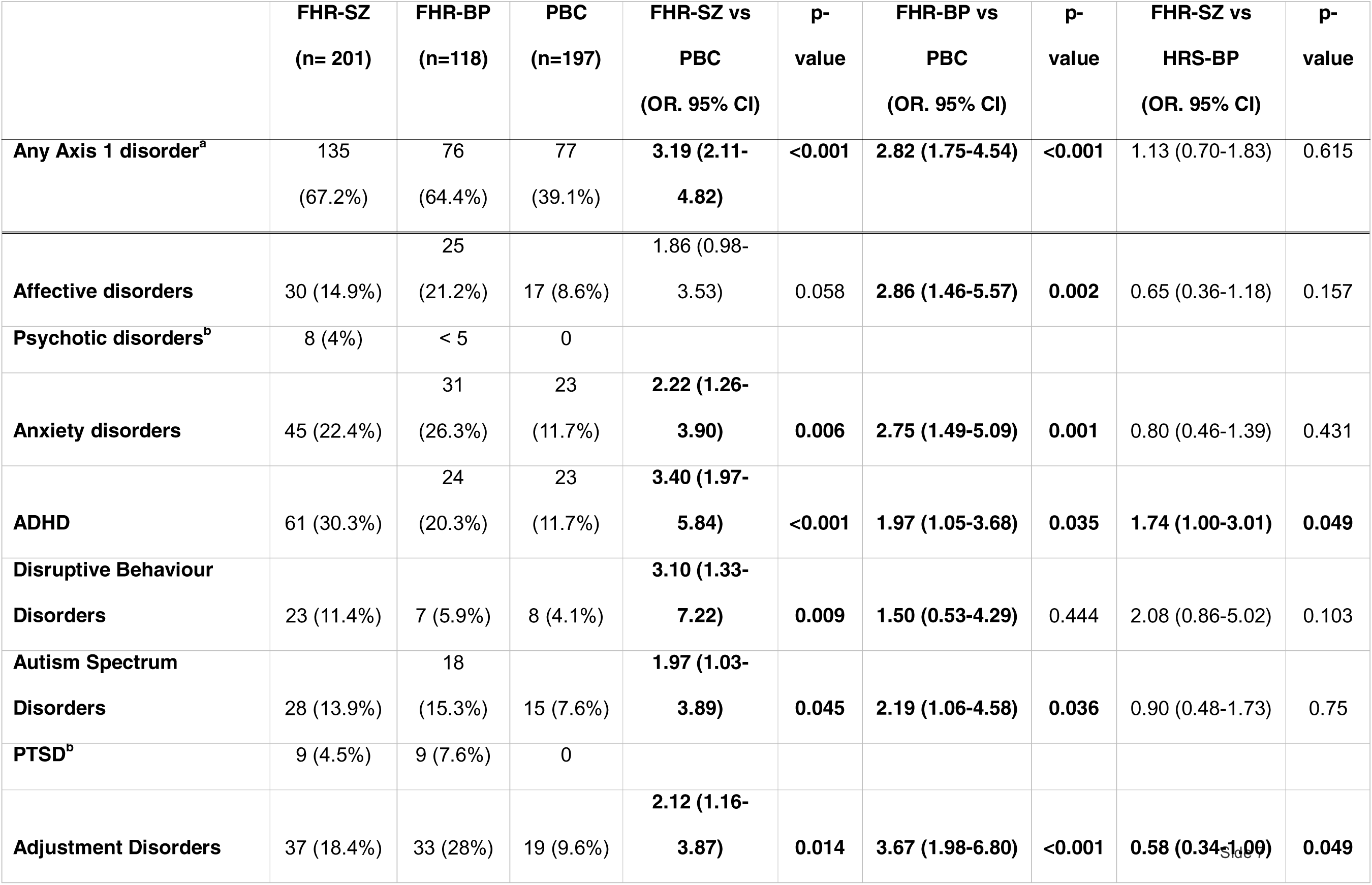
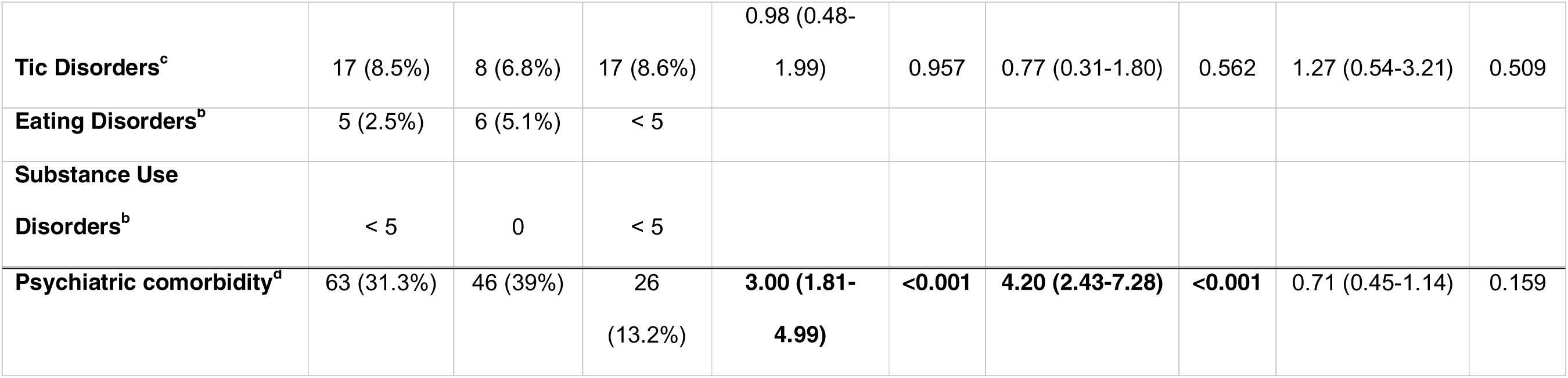
Cumulative incidence of DSM-IV and DSM-V Axis 1 mental disorders and psychiatric comorbidity in 516 15-year-old adolescents in The Danish High Risk and Resilience Study assessed by the K-SADS-PL interview at least once i.e., either at baseline (age 7), four-year follow up (age 11), or eight-year follow-up (age 15). FHR-BP: Adolescents at familial high-risk of bipolar disorder; FHR-SZ: Adolescents at familial high-risk of schizophrenia spectrum disorders; PBC: Population-based controls. K-SADS-PL, Schedule for Affective Disorders and Schizophrenia for School-Age Children Present and Lifetime Version. ^a^ Any Axis I disorder: Elimination disorders, transient and unspecified tics, and specific phobias were excluded; ^b^To few cases to calculate pair-wise comparison; ^c^ Tic disorders includes Tourette’s disorder and chronic tic disorder; ^d^ Psychiatric comorbidity defined as the adolescent has met criteria for two or more different disorder categories at baseline (between ages 0-7), four-year follow-up (between ages 7-11) or follow-up (between ages 11-15).

Both FHR groups exhibited a significantly higher cumulative incidence of ADHD, with threefold higher odds in the group of adolescents at FHR-SZ and nearly twofold higher odds in the group of adolescents at FHR-BP compared with the PBC group (Table 3). Also, the cumulative incidence of ADHD was significantly higher in the group of adolescents at FHR-SZ than in the FHR-BP group.

The cumulative incidence of affective disorders, anxiety disorders and adjustment disorders were approximately threefold higher in the FHR-BP group compared with the PBC group (Table 3). Adolescents at FHR-BP also had the highest cumulative incidence of autism spectrum disorders (15.3%). In the group of adolescents at FHR-SZ the cumulative incidence of anxiety disorders and adjustment disorders was significantly higher compared with PBC. Disruptive behavior disorders were most common in adolescents at FHR-SZ, with threefold higher odds compared with the PBC group (Table 3).

Psychotic disorders and PTSD were observed exclusively in the FHR groups; however, the low number of cases precluded calculation of group differences. Eating disorders occurred across all three groups, with the highest proportions among adolescents at FHR-BP. Substance use disorders were rare, appearing only in the FHR-SZ and PBC groups. As with other outcomes, the few cases precluded meaningful group comparisons (Table 3).

In general, adolescents at FHR-SZ or FHR-BP had higher cumulative incidences across main diagnostic categories from ages 7 and 11 to 15 compared to PBCs. However, development of incidence over time was similar across the three groups (all p-values,>0.05; lowest p = 0.13) (Figure 1 and supplementary figures 1-8).

**Figure 1.**
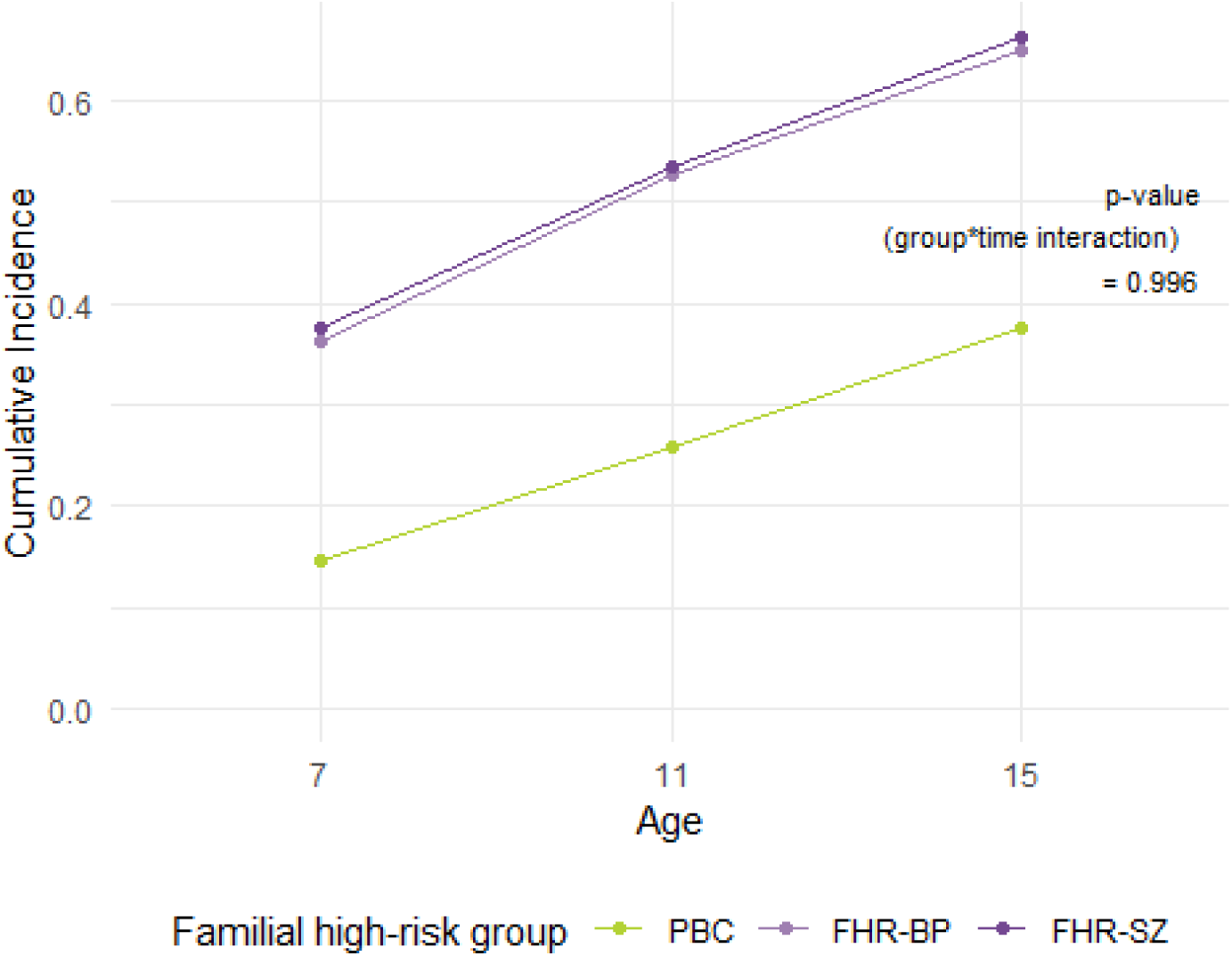
The cumulative incidence of any Axis 1 DSM-IV and DSM-V disorder from baseline (age 7), to four-year follow-up (age 11), and to eight-year follow-up (age 15). FHR-SZ: Adolescents at familial high-risk of schizophrenia; FHR-BP: Adolescents at familial high-risk of bipolar disorder; PBC: Population-based controls. A Wald test is used to test the interaction between time and familial high-risk group.

### Disorder persistency from ages 11-15

A total of 396 participants completed K-SADS-PL interviews both at age 11 and at age 15 (FHR-SZ 147; FHR-BP 94; PBC 155). Among these, 61.3 % of PBC adolescents had no mental disorder at either ages, compared to 42.6% in adolescents at FHR-BP and 34.7% of adolescents in the FHR-SZ group (Figure 2). Persistent disorders were twice as common in adolescents at FHR-SZ and adolescents at FHR-BP compared with those in PBC group. Incident disorders were relatively similar across groups. The distribution of mental disorder status categories differed significantly across the three study groups (χ² (6) = 28.0, p < .001; Figure 2).

**Figure 2.**
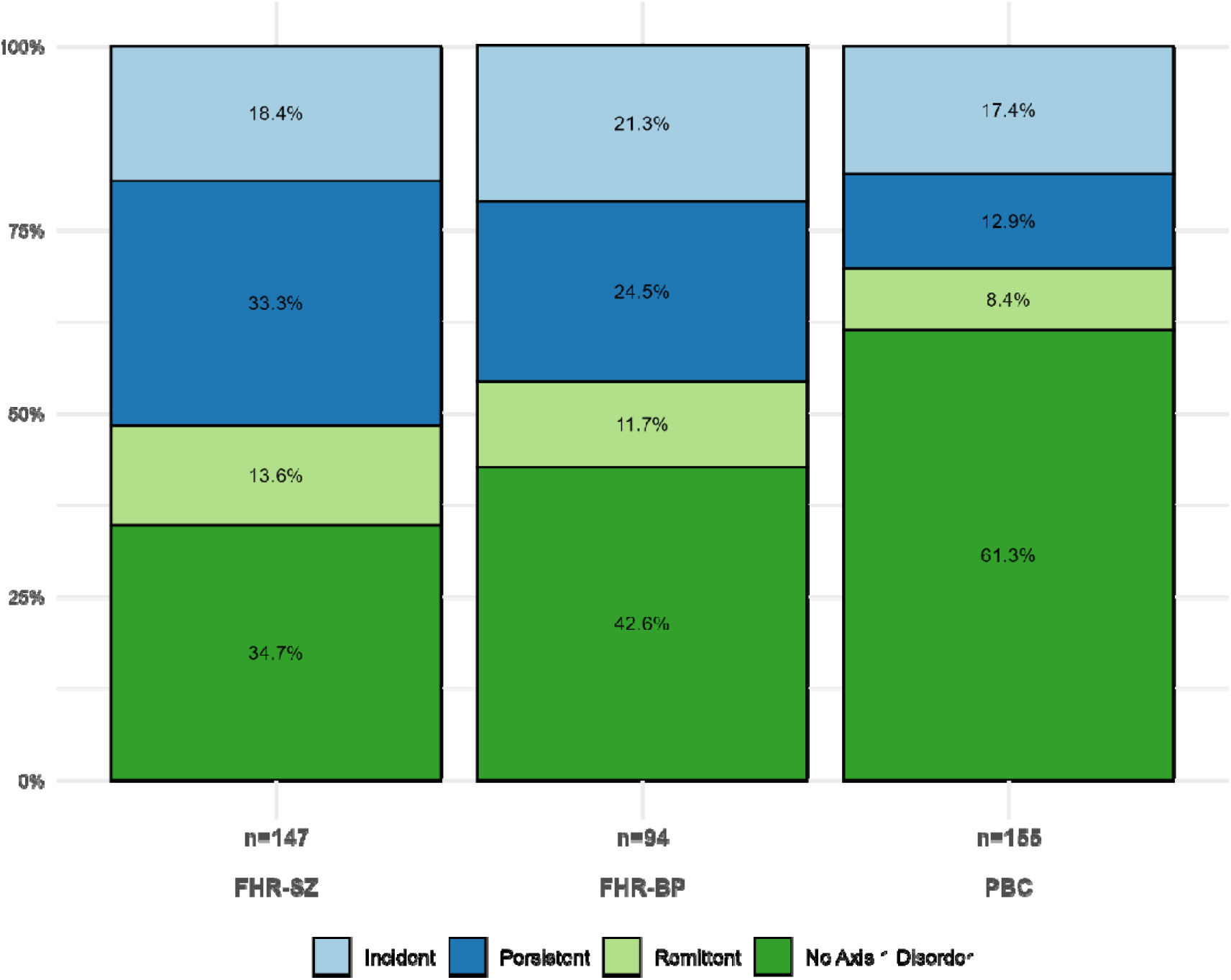
Disorder persistency classification in The Danish High Risk and Resilience Study. Participants with valid K-SADS-PL assessments at age 11 and age 15 were categorized based on changes in mental disorder status (Total N: 396; FHR-SZ 147; FHR-BP 94; PBC 155). FHR-BP: Adolescents at familial high-risk of bipolar disorder; FHR-SZ, Adolescents at familial high-risk of schizophrenia spectrum disorders; PBC, Population-based controls. Incident: Any mental disorder absent at age 11 but present at age 15; Persistent: Any mental disorder present at both ages 11 and 15; Remittent: Any mental disorder present at age 11 but absent at age 15; No disorder: Any mental disorder absent at both ages 11 and 15. Differences in category distribution across all three study groups were tested using a Chi-square test.

### Global functioning

By age 15 (N = 516) estimated GCAS mean values differed significantly among the three groups (FHR-SZ 64.8; FHR-BP 70.1; PBC 76.6, p <.001). Pairwise comparisons indicated significant differences between FHR-SZ and PBC (p <.001), FHR-BP and PBC (p = .012). Regarding the trajectories of CGAS scores from ages 7 to 15, the time-by-group interaction was non-significant (p = .329). Across all three time points, PBC consistently showed the highest scores, FHR-BP intermediate scores, and FHR-SZ the lowest (Figure 3).

**Figure 3:**
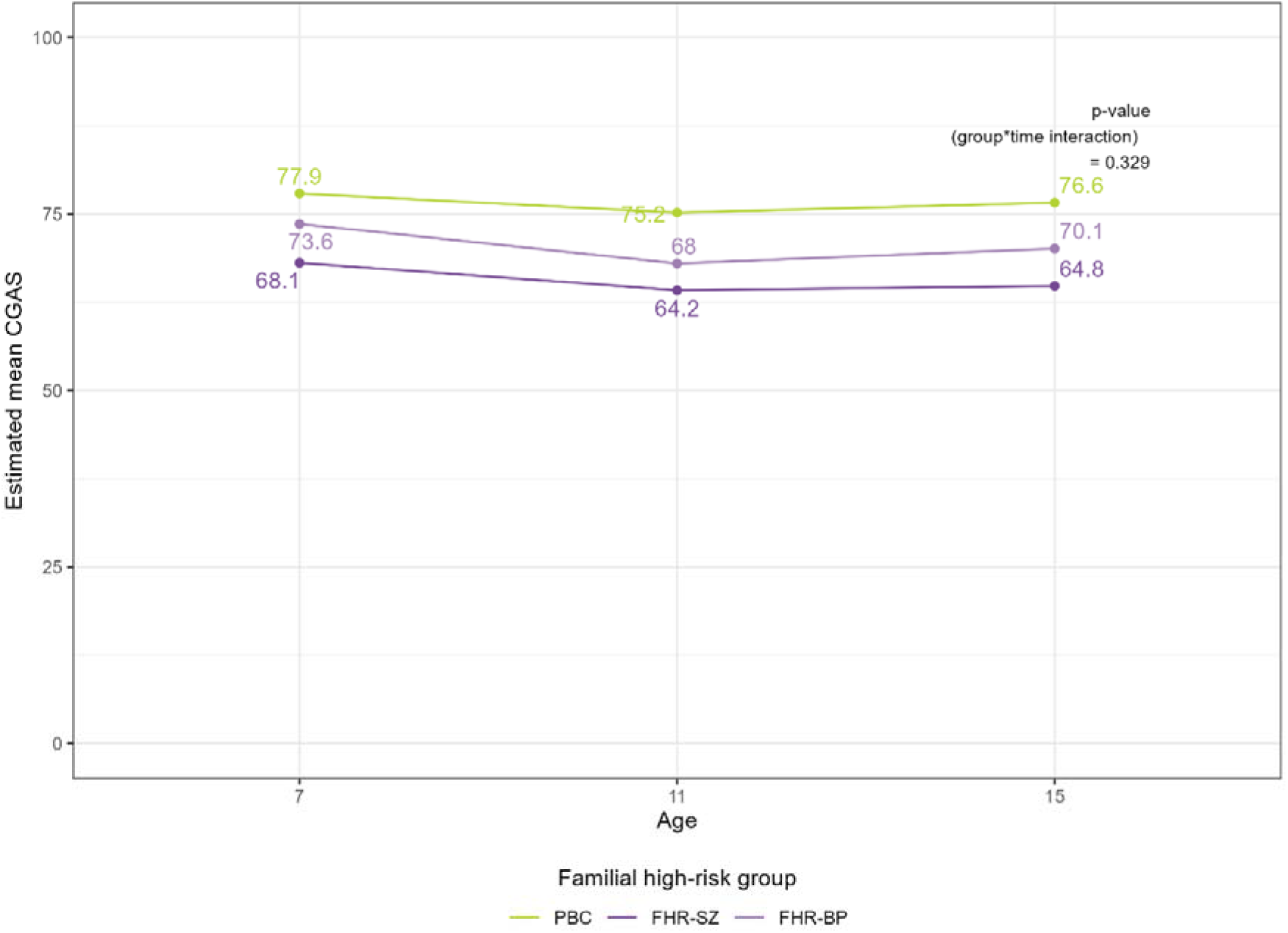
Development of Children’s Global Functioning Scale (CGAS) at baseline (age 7), four-year follow-up (age 11), and eight-year follow-up (age 15) in The Danish High Risk and Resilience Study. FHR-BP: Adolescents at familial high-risk of bipolar disorder; FHR-SZ, Adolescents at familial high-risk of schizophrenia spectrum disorders; PBC, Population-based controls.

## Discussion

In our eight-year follow-up study of FHR 15-year-old adolescents, the four-year prevalence showed a twofold higher risk of any Axis I disorder for FHR-BP and FHR-SZ compared with the PBC group, while the cumulative incidence indicated a threefold higher risk of mental disorders in the FHR groups. Both FHR groups also demonstrated lower levels of global functioning and higher disorder persistency. Between-group differences were non-significant for both mental disorders and global functioning, suggesting that these differences remained stable across ages 7, 11, and 15.

Our findings are consistent with previous results in the Danish High Risk and Resilience study at baseline (age 7) and at the four-year follow-up (age 11)(Ellersgaard et al., 2018; Gregersen et al., 2022). In our study both adolescents at FHR-SZ or FHR-BP displayed higher prevalence and cumulative incidence of ADHD compared to the PBC group with the incidence being highest in the FHR-SZ group. These findings are in line with results from previous FHR studies, providing further evidence for a more pronounced neurodevelopmental illness course, particularly in children at FHR of SZ(De la Serna et al., 2021, 2025; Sanchez-Gistau et al., 2015; Setiaman et al., 2024). Studies have indicated that ADHD in individuals at FHR-SZ may precede the later development of psychosis(Keshavan et al., 2008). In contrast, a review of offspring cohort studies concluded that childhood ADHD is not a reliable antecedent of BP, whereas anxiety appears to be a clear marker of increased risk later in life (Duffy, 2012). However, elevated rates of ADHD have been reported in offspring of parents with psychotic spectrum BP who do not respond to lithium treatment—a subgroup linked to poorer outcomes (Duffy, 2012; Duffy et al., 2014; Hui et al., 2019).

Adjustment disorders and anxiety disorders were more frequent in the FHR-BP group, though both FHR groups had significantly higher cumulative incidences compared to PBC. Notably, psychotic disorders were exclusively observed in the FHR groups, with most cases in the FHR-SZ group (n=8), and the cumulative incidence of affective disorders were highest in the FHR-BP group (21%), suggesting increasing differentiation of parental diagnosis. There were no cases of manic episodes or bipolar disorder, which aligns with existing evidence indicating that most cases of bipolar disorder typically have their onset with a depressive episode, often occurring during adolescence or early adulthood (Duffy, Goodday, Keown-Stoneman, & Grof, 2019; Helmink et al., 2024). As the diagnosis of BP is frequently delayed, adversely impacting the prognosis, it is crucial to closely monitor children of parents with BP who present with affective disorders or anxiety to facilitate the initiation of preventive interventions and ensure timely diagnosis and treatment(Levit, Nunez, Morton, & Keramatian, 2024).

Although our findings may suggest potential developmental trajectories in mental disorders among FHR offspring, the broader picture indicates that the FHR adolescents exhibit a wide range of mental disorders, which do not necessarily mirror those of their parents. Mental disorders in childhood and adolescence are associated with higher risk of developing SZ and BP (Maibing et al., 2015; Meier et al., 2018). However, it remains essential to avoid overinterpreting early diagnoses as definitive predictors of later psychopathology, particularly given that psychiatric diagnoses in childhood and adolescence are often unstable (Krantz et al., 2025).

While the FHR groups consistently showed higher cumulative incidences of mental disorders at all three time points, the change in group differences over time was not statistically significant. This confirms previous findings of stable, early-emerging group differences in psychopathology incidence, with no evidence that the trajectory differed between the FHR and control groups and is consistent with our four-year follow-up study (De la Serna et al., 2021; Gregersen et al., 2022). Whether these patterns will change during late adolescence or early adulthood remains uncertain and may still change as these periods represent peak onset of both SZ and BP (Jauhar, Johnstone, & McKenna, 2022; Nierenberg et al., 2023).

The distribution of disorder status categories (persistent, incident, remittent, no disorder) between ages 11 and 15 differed significantly across the three groups, reflecting a higher proportion of persistent disorder in the FHR groups and a higher proportion of no disorder in the PBC group. These patterns suggest that FHR adolescents were less likely to experience remission, which may partly be explained by higher psychiatric comorbidity in the FHR groups. However, because ADHD and autism were more common in both FHR groups—and these conditions are generally considered to follow a stable course over time—the findings should be interpreted within this context.

Adolescents in both FHR groups demonstrated significantly lower global functioning compared to the PBC group at age 15. This is in line with our previous findings and consistent with the broader literature on functional outcomes among high-risk offspring(Ellersgaard et al., 2018; Gregersen et al., 2022; Helmink et al., 2024). It is likely that our findings reflect the burden of mental disorders within these groups as the CGAS scores take into account whether the adolescent’s daily functioning is affected by symptoms of a mental disorder. This is clinically relevant in the group of adolescents at FHR-SZ, where the mean score difference compared to PBC exceeds 10 points (i.e. 10% of the scale). Consistent with previous research, the lower level of functioning observed in the FHR groups persisted across all assessment points, from ages 7 and 11 through age 15 (De la Serna et al., 2021). These differences should also be interpreted within the broader familial context associated with severe mental illness. In the FHR-SZ and FHR-BP groups, primary caregivers were more likely to experience reduced employment and single parenthood, and caregivers in the FHR-SZ group also had lower educational attainment compared with the PBC group. Thus, these families’ abilities to provide stimulation and inspiration for the adolescents may be impaired, in e.g. not having financial resources to support leisure time activities, trips, cultural activities etc. Such social factors are closely embedded with the course of severe mental illness and resilience and are therefore not clearly separable from psychopathological risk and global functioning captured by measures such as the CGAS.

### Strengths and limitations

This is the first prospective study to compare the psychopathological development of same-aged adolescents at FHR-SZ or FHR-BP with a population-based control group at from childhood to adolescence. Using Danish registers to recruit families represents a key strength of this nationwide cohort. All interviews were conducted face to face by trained healthcare professionals using the validated instrument, K-SADS-PL. Interviewers were blinded to familial high-risk status, thus preventing any bias in symptom assessment and diagnosis. The high retention rates at both four-year (89%) and the present eight-year follow-ups (82%), together with dropout analyses showing no differences between participants and non-participants regarding FHR-status, sex distribution, global functioning, or mental disorder, reduce the likelihood of attrition bias and support the generalizability of our findings.

Several limitations should be acknowledged, too. To assess diagnostic development, we imputed diagnostic information based on diagnostic history for individuals not participating at age 11 and/or age 15.

Consequently, individuals who did not fulfill the criteria for a diagnosis at the latest assessment were still classified as healthy. This approach might have resulted in an underestimation of the cumulative incidence of mental disorders in our study. Although the VIA study did not include any interventions for the participants, study participation itself may have led to increased awareness and knowledge about mental disorders, potentially serving as a preventive intervention, which could have influenced the results. In some cases, it was our legal and ethical obligation to provide either a notification to the municipality or a referral to mental health services to ensure an adolescents’ assessment, treatment or support. Additionally, at baseline of the Danish High Risk and Resilience Study non-participants showed higher socioeconomic and health disadvantages which might indicate a selection bias in the cohort(Krantz et al., 2023). Finally, the FHR-BP group is smaller than the FHR-SZ group and the PBC group, which reduces the statistical power to detect subtle between-group differences.

## Conclusion

Using data from an eight-year follow-up, this nationwide cohort study contributes with novel findings on the four-year prevalence, cumulative incidence and persistency of mental disorders, as well as global functioning in 15-year-old adolescents at FHR-SZ or FHR-BP, compared with PBC. We found that in adolescents with FHR-SZ or FHR-BP, a higher prevalence of mental disorders detectable already in childhood, persisted into adolescence. Moreover, adolescents with FHR-SZ consistently exhibited the lowest levels of functioning, followed by those with FHR-BP, with disparities emerging in early childhood also persisting over time. The increased risk of mental disorders was evident in both FHR groups. Notably, these adolescents present with a broad range of mental disorders, not confined to those for which they have a familial predisposition. The significantly elevated risk of ADHD among adolescents with FHR-SZ compared to those with FHR-BP and PBC could suggest a more pronounced neurodevelopmental impairment, while the higher rates of affective disorders in FHR-BP adolescents may reflect a course more aligned with mood disorders. These findings may help understand the development of mental disorders and the need for preventive interventions for FHR-SZ and FHR-BP individuals. To fully understand the impact of mental disorders during childhood and adolescence, follow-up into adulthood is essential.

## Acknowledgements

We thank all the families who participated in this study for their time and commitment. We acknowledge Sara Nørgard Jensen from Sanos for her statistical expertise. We are grateful to our research coordinator, Tania Storm, for her practical organization of the data collection, to Christina Weise Wittrock for her assistance with interviewer support and practical tasks, and to Hanne Junge Larsen for her invaluable administrative support. Furthermore, we thank Henriette Norman Hansen, Klara Askær Thylin, Marie Nymand, and Nanna Lawaetz for their contributions to data collection. The VIA 15 study was funded by Lundbeckfonden, the Novo Nordisk Foundation, and the Research Fund of the Mental Health Services in the Capital Region of Denmark

## Ethics Statement

The Danish Data Protection Agency approved the VIA 7, VIA 11, and VIA 15 studies (approval number: P-2019-273. The Danish Committee on Health Research Ethics concluded that ethical approval was unnecessary due to the study’s observational nature for the VIA 7 study, while ethical approval was obtained for the VIA 11 study (Protocol number: H-16043682, approved January 2, 2017) and the VIA 15 study (Protocol number: H-20067908, approved March 24, 2021). Following a thorough explanation of the procedures, the legal guardians provided written informed consent for their own and their child’s participation, while adolescents provided oral assent.

## Data Availability

First author, Doris Helena Bjarnadóttir Streymá had full access to all the data in the study and takes responsibility for the integrity of the data and the accuracy of the data analysis. Participants’ identities were protected by assigning unique identification codes, and all study data were securely stored in accordance with ethical standards. The data collected and analyzed in this study are subject to ethical restrictions and therefore cannot be made publicly available. However, data may be made available upon reasonable request to the corresponding author, Doris Helena Bjarnadóttir Streymá.

## References

Anttila, V., Bulik-Sullivan, B., Finucane, H. K., Walters, R. K., Bras, J., Duncan, L., Escott-Price, V., et al. (2018). Analysis of shared heritability in common disorders of the brain. Science, 360(6395), 8757. American Association for the Advancement of Science.

Davidsen, K. A., Munk-Laursen, T., Foli-Andersen, P., Ranning, A., Harder, S., Nordentoft, M., & Thorup, A. A. E. (2022). Mental and pediatric disorders among children 0–6 years of parents with severe mental illness. Acta Psychiatrica Scandinavica, 145(3), 244–254. John Wiley and Sons Inc.

De la Serna, E., Ilzarbe, D., Sugranyes, G., Baeza, I., Moreno, D., Rodríguez-Toscano, E., Espliego, A., et al. (2021). Lifetime psychopathology in child and adolescent offspring of parents diagnosed with schizophrenia or bipolar disorder: A 2-year follow-up study. European Child and Adolescent Psychiatry, 30(1), 117–129. Springer Science and Business Media Deutschland GmbH.

De la Serna, E., Moreno, D., Sugranyes, G., Camprodon-Boadas, P., Ilzarbe, D., Bigorra, A., Mora-Maltas, B., et al. (2025). Effects of parental characteristics on the risk of psychopathology in offspring: A 4-year follow-up study. European Child & Adolescent Psychiatry 2025 34:10, 34(10), 3035–3045. Springer.

Duffy, A. (2012). The nature of the association between childhood ADHD and the development of bipolar disorder: A review of prospective high-risk studies. American Journal of Psychiatry, 169(12), 1247– 1255. American Psychiatric Association.

Duffy, A., Goodday, S., Keown-Stoneman, C., & Grof, P. (2019). The emergent course of bipolar disorder: Observations over two decades from the Canadian high-risk offspring cohort. American Journal of Psychiatry, 176(9), 720–729. American Psychiatric Association.

Duffy, A., Horrocks, J., Doucette, S., Keown-Stoneman, C., McCloskey, S., & Grof, P. (2014). The developmental trajectory of bipolar disorder. The British Journal of Psychiatry, 204(2), 122–128. Cambridge University Press.

Ellersgaard, D., Jessica Plessen, K., Richardt Jepsen, J., Soeborg Spang, K., Hemager, N., Klee Burton, B., Jerlang Christiani, C., et al. (2018). Psychopathology in 7-year-old children with familial high risk of developing schizophrenia spectrum psychosis or bipolar disorder – The Danish High Risk and Resilience Study—VIA 7, a population-based cohort study. World Psychiatry, 17(2), 210–219. Blackwell Publishing Ltd.

Erlenmeyer-Kimling & Cornblatt. (1987). The New York High-Risk Project: A Followup Report. Schizophrenia Bulletin, 13(3), 451–461.

Gregersen, M., Søndergaard, A., Brandt, J. M., Ellersgaard, D., Rohd, S. B., Hjorthøj, C., Ohland, J., et al. (2022). Mental disorders in preadolescent children at familial high-risk of schizophrenia or bipolar disorder-a four-year follow-up study The Danish High Risk and Resilience Study, VIA 11. Journal of Child Psychology and Psychiatry, 63(9), 1046–1056.

Hans, Auerbach, J., Styr, B., & Marcus, J. (2004). Offspring of Parents With Schizophrenia: Mental Disorders During Childhood and Adolescence. Schizophrenia Bulletin, 30(2), 303–315.

Helmink, F. G. L., Mesman, E., & Hillegers, M. H. J. (2024). Beyond the Window of Risk? The Dutch Bipolar Offspring Study: 22-Year Follow-up. Retrieved January 3, 2025, from www.jaacap.org

Hui, T. P., Kandola, A., Shen, L., Lewis, G., Osborn, D. P. J., Geddes, J. R., & Hayes, J. F. (2019). A systematic review and meta-analysis of clinical predictors of lithium response in bipolar disorder. Acta Psychiatrica Scandinavica, 140(2), 94–115. John Wiley & Sons, Ltd.

Jauhar, S., Johnstone, M., & McKenna, P. J. (2022). Schizophrenia. The Lancet, 399(10323), 473–486. Elsevier B.V.

Kaufman, J., Birmaher, B., Brent, D., Rao, U., Flynn, C., Moreci, P., Williamson, D., et al. (1997). Schedule for affective disorders and schizophrenia for school-age children-present and lifetime version (K-SADS-PL): Initial reliability and validity data. Journal of the American Academy of Child and Adolescent Psychiatry, 36(7), 980–988. Elsevier Inc.

Keshavan, M., Montrose, D. M., Rajarethinam, R., Diwadkar, V., Prasad, K., & Sweeney, J. A. (2008). Psychopathology among offspring of parents with schizophrenia: Relationship to premorbid impairments. Schizophrenia Research, 103(1–3), 114–120.

Krantz, M. F., Dalsgaard, S., Osler, M., Jorgensen, M. B., Jorgensen, A., & Jørgensen, T. S. H. (2025). Diagnostic trajectories and stability of mental disorders in childhood and adolescence – A nation-wide cohort study using sequence analysis. European Psychiatry, 68(1). Royal College of Psychiatrists.

Krantz, M. F., Hjorthøj, C., Ellersgaard, D., Hemager, N., Christiani, C., Spang, K. S., Burton, B. K., et al. (2023). Examining selection bias in a population-based cohort study of 522 children with familial high risk of schizophrenia or bipolar disorder, and controls: The Danish High Risk and Resilience Study VIA 7. Social Psychiatry and Psychiatric Epidemiology, 58(1), 113–140. Springer Science and Business Media Deutschland GmbH.

Levit, A., Nunez, J.-J., Morton, E., & Keramatian, K. (2024). Factors influencing delays in the diagnosis and treatment of bipolar disorder in adolescents and young adults: A systematic scoping review. European Psychiatry, 67(S1), S297–S297. Cambridge University Press.

Maibing, C. F., Pedersen, C. B., Benros, M. E., Mortensen, P. B., Dalsgaard, S., & Nordentoft, M. (2015). Risk of Schizophrenia Increases After All Child and Adolescent Psychiatric Disorders: A Nationwide Study. Schizophrenia Bulletin, 41(4), 963–970. Oxford Academic.

Maughan, B., & Collishaw, S. (2015). Development and psychopathology: A life course perspective. Rutter’s Child and Adolescent Psychiatry: Sixth Edition, 1–16. John Wiley and Sons Ltd.

Maziade, Gingras, Rouleau, Poulin, Jomphe, Paradis, Mérette, et al. (2008). Clinical diagnoses in young offspring from eastern Quebec multigenerational families densely affected by schizophrenia or bipolar disorder. Acta Psychiatrica Scandinavica, 117(2), 118–126.

Meier, S. M., Pavlova, B., Dalsgaard, S., Nordentoft, M., Mors, O., Mortensen, P. B., & Uher, R. (2018). Attention-deficit hyperactivity disorder and anxiety disorders as precursors of bipolar disorder onset in adulthood. The British Journal of Psychiatry, 213(3), 555–560. Cambridge University Press.

Mesman, E., Nolen, W. A., Reichart, C. G., Wals, M., & Hillegers, M. H. J. (2013). The dutch bipolar offspring study: 12-year follow-up. American Journal of Psychiatry, 170(5), 542–549. American Psychiatric Association.

Morosini, P. L., Magliano, L., Brambilla, L., Ugolini, S., & Pioli, R. (2000). Development, reliability and acceptability of a new version of the DSM-IV Social Occupational Functioning Assessment Scale (SOFAS) to assess routine social functioning. Acta Psychiatrica Scandinavica, 101(4), 323–329.

Mors, O., Perto, G. P., & Mortensen, P. B. (2011). The Danish psychiatric central research register. Scandinavian Journal of Public Health, 39(7), 54–57.

Nierenberg, A. A., Agustini, B., Köhler-Forsberg, O., Cusin, C., Katz, D., Sylvia, L. G., Peters, A., et al. (2023). Diagnosis and Treatment of Bipolar Disorder: A Review. JAMA, 330(14), 1370–1380. American Medical Association.

Pedersen, C. B. (2011). The Danish civil registration system. Scandinavian Journal of Public Health, 39(7), 22–25.

Reynolds, C. R., & Kamphaus, R. W. (2003). Reynolds Intellectual Assessment Scales (RIAS). Lutz, FL: Psychological Assessment Resources.

Robinson, N., & Bergen, S. E. (2021). Environmental Risk Factors for Schizophrenia and Bipolar Disorder and Their Relationship to Genetic Risk: Current Knowledge and Future Directions. Frontiers in Genetics, 12. Frontiers Media S.A.

Robinson, N., Ploner, A., Leone, M., Lichtenstein, P., Kendler, K. S., & Bergen, S. E. (2024). Environmental risk factors for schizophrenia and bipolar disorder from childhood to diagnosis: A Swedish nested case-control study. Psychological Medicine. Cambridge University Press.

Ross, R. G., & Compagnon, N. (2001). Diagnosis and treatment of psychiatric disorders in children with a schizophrenic parent. Schizophrenia Research, 50(1–2), 121–129. Elsevier.

Sanchez-Gistau, V., Romero, S., Moreno, D., de la Serna, E., Baeza, I., Sugranyes, G., Moreno, C., et al. (2015). Psychiatric disorders in child and adolescent offspring of patients with schizophrenia and bipolar disorder: A controlled study. Schizophrenia Research, 168(1–2), 197–203. Elsevier.

Setiaman, N., Mesman, E., van Haren, N., & Hillegers, M. (2024). Emerging psychopathology and clinical staging in adolescent offspring of parents with bipolar disorder or schizophrenia—A longitudinal study. Bipolar Disorders, 26(1), 58–70. John Wiley and Sons Inc.

Shaffer, D., Gould, M. S., Brasic, J., Fisher, P., Aluwahlia, S., & Bird, H. (1983). A Children’s Global Assessment Scale (CGAS). Archives of General Psychiatry, 40(11), 1228–1231. American Medical Association.

Shah, J. L., Tandon, N., Montrose, D. M., Mermon, D., Eack, S. M., Miewald, J., & Keshavan, M. S. (2019). Clinical psychopathology in youth at familial high risk for psychosis. Early Intervention in Psychiatry, 13(2), 297–303. Blackwell Publishing.

Solmi, M., Radua, J., Olivola, M., Croce, E., Soardo, L., Salazar de Pablo, G., Il Shin, J., et al. (2022). Age at onset of mental disorders worldwide: Large-scale meta-analysis of 192 epidemiological studies. Molecular Psychiatry, 27(1), 281–295. Springer Nature.

Thorup, A. A. E., Hemager, N., Bliksted, V. F., Greve, A. N., Ohland, J., Wilms, M., Rohd, S. B., et al. (2022). The Danish High-Risk and Resilience Study—VIA 15 – A Study Protocol for the Third Clinical Assessment of a Cohort of 522 Children Born to Parents Diagnosed With Schizophrenia or Bipolar Disorder and Population-Based Controls. Frontiers in Psychiatry, 13, 809807. Frontiers Media S.A.

Thorup, A. A. E., Hemager, N., Søndergaard, A., Gregersen, M., Prøsch, Å. K., Krantz, M. F., Brandt, J. M., et al. (2018). The Danish High Risk and Resilience Study-VIA 11: Study Protocol for the First Follow-Up of the VIA 7 Cohort -522 Children Born to Parents With Schizophrenia Spectrum Disorders or Bipolar Disorder and Controls Being Re-examined for the First Time at Age 11. Frontiers in psychiatry, 9, 661. Frontiers Media S.A.

Thorup, A. A. E., Jepsen, J. R., Ellersgaard, D. V., Burton, B. K., Christiani, C. J., Hemager, N., Skjærbæk, M., et al. (2015). The Danish High Risk and Resilience Study—VIA 7—A cohort study of 520 7-year-old children born of parents diagnosed with either schizophrenia, bipolar disorder or neither of these two mental disorders. BMC Psychiatry, 15(1), 1–15. BioMed Central Ltd.

Uher, R., Pavlova, B., Radua, J., Provenzani, U., Najafi, S., Fortea, L., Ortuño, M., et al. (2023). Transdiagnostic risk of mental disorders in offspring of affected parents: A meta-analysis of family high-risk and registry studies. World Psychiatry, 22(3), 433–448. John Wiley and Sons Inc.

