## Supplementary Material for "Mental disorders in adolescents at familial high-risk of schizophrenia or bipolar disorder and population-based controls – an eight-year follow-up study, The Danish High Risk and Resilience Study, VIA 15"

Supplementary Table 1. Dropout analyses comparing familial high-risk group, sex, global functioning and lifetime history of any mental disorder measured at age 7 years for adolescents participating vs not participating in K-SADS-PL interview at age 15.

|  | Not participating in K-SADS-PL at age 15 (N=106) | Participating in K-SADS-PL at age 15 (N=416) | P-value |
| --- | --- | --- | --- |
| Familial high-risk group, n (%) |  |  |  |
| Schizophrenia | 48 (45.3%) | 154 (37.0 %) | **0.656** |
| Bipolar disorder | 22 (20.8 %) | 98 (23.6 %) |  |
| Population based controls | 36 (34.0 %) | 164 (39.4 %) |  |
| Females, N (%) | 43 (40.6 %) | 199 (47.8%) | **0.407** |
| CGAS, mean (SD)* | 70.2 (16.2) | 73.7 (14.8) | **0.114** |
| Any axis 1 diagnosis, N (%) * | 33 (33.0 %) | 114 (37.4 %) | **0.336** |

Drop-out analysis tested for differences in the VIA7 data between participants included vs. non-included in the present study. Two-tailed t-tests were used to assess CGAS. A chi-square test assessed familial high-risk group, sex and prevalence of lifetime axis-I disorders. Significant p-values are shown in bold. Abbreviations: CGAS: Children’s Global Assessment Scale; SD: standard deviation. *Missing data in VIA7: CGAS (n=8), axis-I disorders (n=8).

Supplementary Figure 1. The cumulative incidence of DSM-IV and DSM-V Affective disorders from baseline (age 7), to four-year follow-up (age 11), and to eight-year follow-up (age 15). FHR-SZ: Adolescents at familial high-risk of schizophrenia; FHR-BP: Adolescents at familial high-risk of bipolar disorder; PBC: Population-based controls. A Wald test is used to test the interaction between time and familial high-risk group.

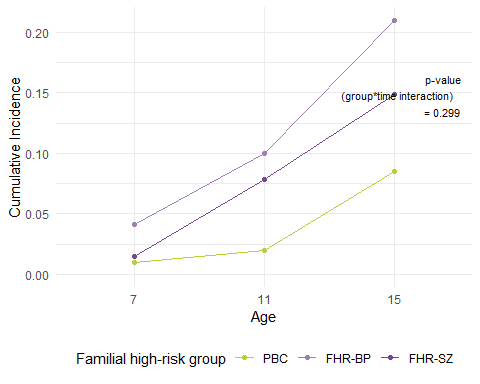

Supplementary Figure 2. The cumulative incidence of DSM-IV and DSM-V Anxiety disorders from baseline (age 7), to four-year follow-up (age 11), and to eight-year follow-up (age 15). FHR-SZ: Adolescents at familial high-risk of schizophrenia; FHR-BP: Adolescents at familial high-risk of bipolar disorder; PBC: Population-based controls. A Wald test is used to test the interaction between time and familial high-risk group.

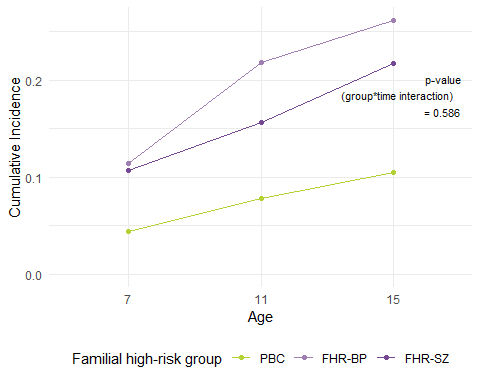

Supplementary Figure 3. The cumulative incidence of DSM-IV and DSM-V Disruptive Behaviour disorders from baseline (age 7), to four-year follow-up (age 11), and to eight-year follow-up (age 15). FHR-SZ: Adolescents at familial high-risk of schizophrenia; FHR-BP: Adolescents at familial high-risk of bipolar disorder; PBC: Population-based controls. A Wald test is used to test the interaction between time and familial high-risk group.

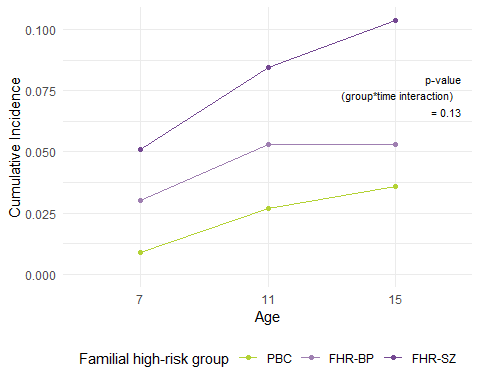

Supplementary Figure 4. The cumulative incidence of DSM-IV and DSM-V ADHD from baseline (age 7), to four-year follow-up (age 11), and to eight-year follow-up (age 15). FHR-SZ: Adolescents at familial high-risk of schizophrenia; FHR-BP: Adolescents at familial high-risk of bipolar disorder; PBC: Population-based controls. A Wald test is used to test the interaction between time and familial high-risk group.

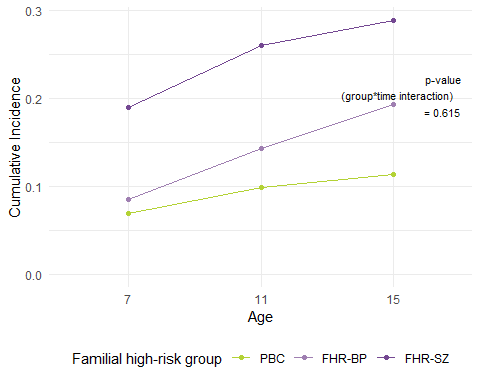

Supplementary Figure 5. The cumulative incidence of DSM-IV and DSM-V Autism Spectrum Disorders from baseline (age 7), to four-year follow-up (age 11), and to eight-year follow-up (age 15). FHR-SZ: Adolescents at familial high-risk of schizophrenia; FHR-BP: Adolescents at familial high-risk of bipolar disorder; PBC: Population-based controls. A Wald test is used to test the interaction between time and familial high-risk group.

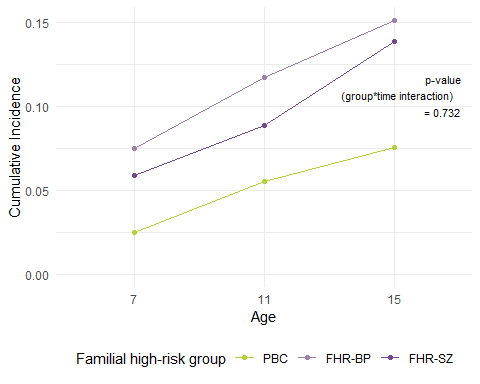

Supplementary Figure 6. The cumulative incidence of DSM-IV and DSM-V Adjustment Disorders from baseline (age 7), to four-year follow-up (age 11), and to eight-year follow-up (age 15). FHR-SZ: Adolescents at familial high-risk of schizophrenia; FHR-BP: Adolescents at familial high-risk of bipolar disorder; PBC: Population-based controls. A Wald test is used to test the interaction between time and familial high-risk group.

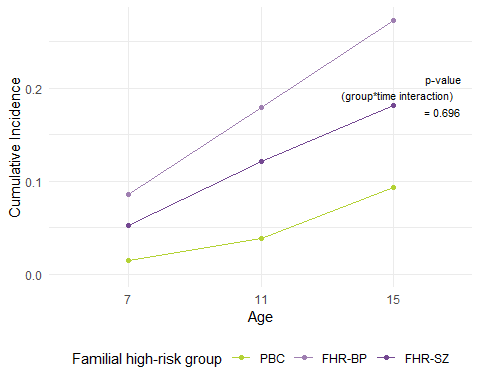

Supplementary Figure 7. The cumulative incidence of DSM-IV and DSM-V Tic Disorders from baseline (age 7), to four-year follow-up (age 11), and to eight-year follow-up (age 15). FHR-SZ: Adolescents at familial high-risk of schizophrenia; FHR-BP: Adolescents at familial high-risk of bipolar disorder; PBC: Population-based controls. A Wald test is used to test the interaction between time and familialhigh-risk group.

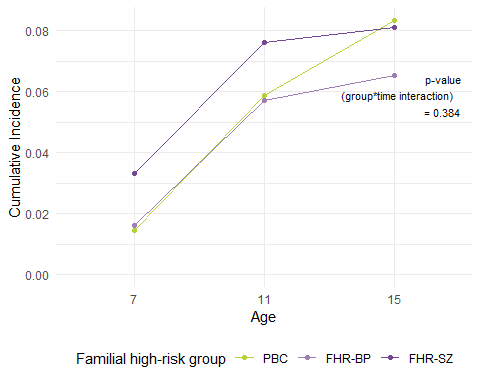

Supplementary Figure 8. The cumulative incidence of two or more DSM-IV and DSM-V disorders from baseline (age 7), to four-year follow-up (age 11), and to eight-year follow-up (age 15). FHR-SZ: Adolescents at familial high-risk of schizophrenia; FHR-BP: Adolescents at familial high-risk of bipolar disorder; PBC: Population-based controls. A Wald test is used to test the interaction between time and familial high-risk group.

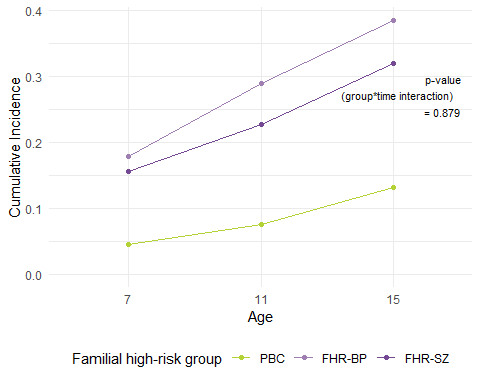
